# The Impact of Mass Exodus on the Resurgence of COVID19 Cases: Study Case of Regions in Indonesia

**DOI:** 10.1101/2021.12.06.21267391

**Authors:** Nuning Nuraini, Kamal Khairudin Sukandar, Wirdatul Aini

## Abstract

The inclusion of the human mobility aspect is essential for understanding the behavior of COVID-19 spread, especially when millions of people travel across borders near Eid Al-Fitr. This study aims at grasping the effect of mass exodus among regions on the active cases of COVID-19 in a mathematical perspective. We construct a multi-region SIQRD (Susceptible-Infected-Quarantined-Recovered-Death) model that accommodates the direct transfer of people from one region to others. The mobility rate is estimated using the proposed Dawson-like function, which requires the Origin-Destination Matrix data. Assuming only susceptible, unapparent infected, and recovered individuals travel near Eid Al-Fitr, the rendered model is well-depicting the actual data at that time, giving either a significant spike or decline in the number of active cases due to the mass exodus. Most agglomerated regions like Jakarta and Depok City experienced the fall of active cases number, both in actual data and the simulated model. However, most rural areas experienced the opposite, like Bandung District and Cimahi City. This study should confirm that most travelers originated from big cities to the rural regions and scientifically justifies that massive mobility affects the COVID-19 transmission among areas.

## 1 Introduction

The COVID-19 pandemic at the beginning of 2020 echoed in all countries. That high-spreader virus has made leaders in almost all countries address similar policies: restricting the movement of people except for groceries. This policy is expected to reduce human-to-human interactions, leading to the decline of the COVID-19 cases number [1]. This prohibition lasted for most of 2020, even though some relaxation had occurred in some countries. In India, the government eased the lockdown after the significant decline in COVID-19 cases, allowing more people to be mobile. The surge of COVID-19 cases is identified after the first relaxation, which was believed as a direct implication of the relaxation [2].

The Centers for Disease Control and Prevention (CDC) consistently provides recommendations regarding restrictions on the movement of people to suppress the spread of the virus in the population as of March 8th, 2021 [3]. These restrictions on movement are basically based on several factors: the number of COVID-19 cases in the community, exposure during travel, the level of crowds in a place, and so on. It is clear that the higher the number of COVID-19 cases in a community and the more crowded a place is, the greater the chance of being exposed to the virus. This is what provides a strong reason for restricting movement in suppressing the spread of infection.

In line with what the CDC recommends, movement restrictions have also been implemented in several provinces of Indonesia with a high number of COVID-19 cases. For example, Jakarta with the terminology of PSBB (Large-Scale Social Restrictions), which literally limits the movement of Jakarta residents through restrictions on schools and workplaces, cessation of religious activities in places of worship, restrictions on activities in public places or facilities, restrictions on socio-cultural activities, to transportation activities. Until the end of 2020, the PSBB has been implemented 12 times in Jakarta [4]. Until the end of February 2021, The Macro and Micro PPKM (Enforcement of Community Activity Restrictions) is implemented to respond to a significant increase in cases in Jakarta, reaching more than 3,000 new cases per day [5]. In general, PPKM is similar to PSBB but applies to the Java and Bali regions.

Ahead of Eid Al-Fitr 1442 Hijri, a mass exodus (a trip to one’s hometown) is unavoidable which will lead millions of people leaving their odyssey [6]. Even though there were a significant drop in the number of travelers in 2020, it is expected that its number will back increasing in 2021 [7]. The massive mobility is believed to trigger more infections since unapparent infected could travel across borders. Depicted earlier in China, the human mobility due to the Chinese New Year does affect the variations in disease incidence between cities. Cities that are closer to the epidemic center and with more population in the urban area will face higher risks of disease incidence [8].

In this study, the multi-region mathematical model is constructed which accommodates the mobility of people among regions. The model generalizes the SIQRD model which split the population into five groups: susceptible, infected, quarantined, recovered, and deaths. The additional parameters representing the time-dependent mobility rate from one region to another is depicted by the modified Dawson Function. The such parameter is estimated by means of the proposed algorithm requiring the data of Origin-Destination matrix. The numerical simulations of the constructed model depict how the mobility rate between regions can affect the surge or decline in the number of COVID-19 cases.

## 2 Context

### 2.1 *Mudik*: Indonesia’s Annual Mass Exodus

*Mudik* or *Balik Kampung* is a common terminology commonly used by Indonesian people to represent the activity where migrants or migrant workers return to their hometown or village toward significant holidays, such as Lebaran (Eid al-Fitr) or Chinese New Year [9]. The primary motivation of this tradition is to visit family, especially parents. Further, not few of them return to their home to attend a rare opportunity: a gathering of members of the extended family, which are seldomly seen and typically scattered in other cities, other provinces, or even overseas. Mudik Eid al-Fitr or similar traditions can also be found in other Muslim-majority countries, such as Egypt, and Bangladesh [10].

The average number of total travellers in this exodus hype, especially during Eid al-Fitr, in Indonesia is approximately 30 million [11]. Even though the mudik homecoming travel before Lebaran takes place in most Indonesian urban centers, the highlight is on the nation’s largest urban agglomeration; Greater Jakarta, as millions of Jakartans exit the city by various means of transportation, overwhelming train stations and airports, and also clogging highways. Annually, the figure of travelers during the exodus shows a constant pattern: gradually increasing started from D-14 towards the holy day and reaching its spike two days before the D-Day. The reversed mobility pattern is also shown days after the holiday, representing the return of migrants from their destination [12].

Every year, the total number of Eid al-Fitr homecomings experienced a significant increase except in 2020 [13], as the figure of travellers enormously dropped to almost zero number given the global pandemic. However, based on the survey data retrieved by Angkutan Lebaran Research Group, the expected number of travellers on the 2021 Eid al-Fitr Mudik will increase and thus could increase the chance of infections of COVID-19 due to its increasing rate of people-to-people interaction. Even worse, Jakarta is believed to contain more than 50% of the total recorded cases in Indonesia by April 2021, making the mudik exodus will trigger more virus spreaders to travel [14]. When the Delta variant is combined with human mobility and an uncontrollable situation, it becomes a suitable environment for viruses to spread and mutate [15].

One of the favourite provinces as the *mudik* destination is West Java [16], a neighboring province situated in the southeast of Jakarta. As a home of 48.6 million inhabitants, West Java comprises 27 cities/districts with current active cases more than 30 thousand by April 2021 [17], far below the cases recorded in the Capital City of Jakarta. Every year, the exodus can move more than 20% of the total population of Jakarta into West Java, entering various cities and districts [18]. The massive movement of people from Red Zone Jakarta to West Java is believed to affect the spread of COVID-19 recorded in regions of West Java.

### 2.2 Available Datasets

Public Health Office of West Java provides the data of COVID-19 spread for each city/district, which is made of three time-series data: the number of active cases, recovered, and deceased individuals due to COVID-19 infections [17]. The data interval ranges from early March 2020 until October 2021. In this case study, the data until May 2021 only be considered representing the spread until the time of the mass exodus, Eid al-Fitr on May 13th, 2021. The data used to estimate the parameters in model (1) will be explained further in the next section.

Further, the survey of the Mudik preference of the Indonesian citizen was provided. In a nutshell, the survey collects the preference of the citizens whether they are going to conduct Mudik or not. If yes, the origin and destination regions should be recorded [19]. The number of participants joining this survey is approximately 62,000, distributed more to the Java citizens. The survey data is expected to estimate the origin-destination matrix for the Eid al-Fitr 2021 exodus. The Origin-Destination matrix is a two-dimensional matrix that contains information about the magnitude of the movement between location (zone) within a certain area [20].

## 3 Mathematical Modeling

### 3.1 Generic *SIQRD* Model

The compartment-approach model is based on the generic model first introduced by Kermack and McKendrick [21], but modifying it by accommodating the vaccination, quarantine, and reinfection aspects. Thousands of papers related to the spread of the Covid-19 model involving several aspects have been published. Several papers using data in Indonesia, i.e discuss a simple model with data on the city of Jakarta at the beginning of the pandemic using the Richard curve [22], [23] the SIR model [24], the super spreading problem with Batam - Depok data [25], PSBB in Jakarta [26], scenarios of vaccine implementation in Indonesia [27] and how the model is applied to policymakers [28]. The term stands for susceptible, infected, quarantined, recovered, and deceased, classifying people into five groups. The detailed information of each compartment is given in Table 1.

**Table 1.** Descriptions of variables and parameters in the {*SIQRD*}_*k*_ model.

| Notation | Description | Unit | Value |
| --- | --- | --- | --- |
| $S_k(t)$ | Number of susceptible individuals at time $t$ in region $k$ | individual | - |
| $I_k(t)$ | Number of infected individuals at time $t$ in region $k$ | individual | - |
| $Q_k(t)$ | Number of quarantined individuals at time $t$ and region $k$ | individual | - |
| $R_k(t)$ | Number of recovered individuals at time $t$ and region $k$ | individual | - |
| $D_k(t)$ | Number of deceased cases at time $t$ and region $k$ | individual | - |
| $\beta_k$ | Transmission rate of COVID-19 in region $k$ | 1/day | estimated |
| $\gamma_k$ | Recovery rate due to COVID-19 infections in region $k$ | 1/day | estimated |
| $\delta_k$ | Death rate due to COVID-19 infections in region $k$ | 1/day | estimated |
| $\alpha_{jk}(t)$ | Daily mobility rate from region $j$ to $k$ due to the mass exodus | 1/day | estimated |
| $v_k$ | Vaccination rate conducted in region $k$ | 1/day | estimated |
| $\zeta$ | Reinfection rate of COVID-19 | 1/day | assumed |
| $q$ | Quarantine rate | 1/day | assumed |
| $\eta$ | Vaccine efficacy | - | assumed |
| $\pi$ | Natural recruitment rate | 1/day | assumed |
| $\mu$ | Natural death rate | 1/day | assumed |

Susceptible individuals might be infected after having contact with infected individuals, which resulted in a portion of them moving to compartment *I*. The quarantine rate governs how many of the infected individuals are to be quarantined (hospitalization or self-isolation) and hence lessens the chance of infecting others. Also, vaccine inoculation is simply accommodated by allowing people from the S compartment to move directly to the R compartment after developing the virus-immune system. This modification reduces the number of susceptible ones depending on the vaccine rate and efficacy, which conforms to the primary goal of the vaccination program [29]. Besides, considering the temporary immunity developed after being inoculated or infected by the disease [30], the modification also covers the case in which recovered/vaccinated individuals are being reinfected by the virus, allowing people in R to move back to S, which can be seen in Figure 1.

**Figure 1.**
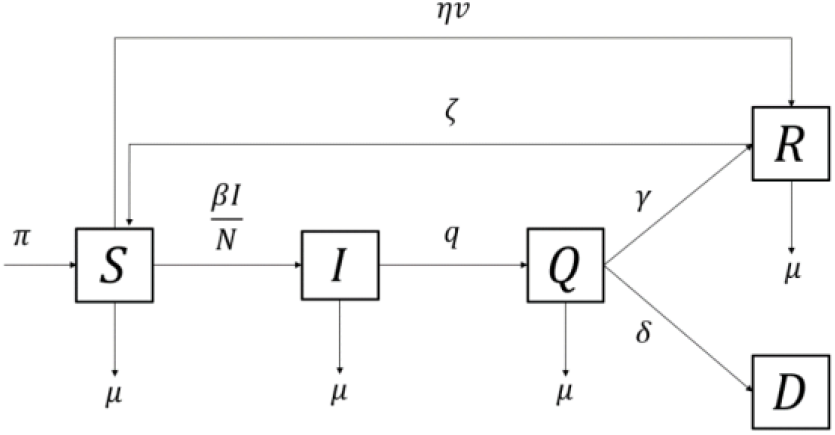
Diagram flow chart of *SIQRD* model.

The mathematical system of *SIQRD* model is given by:

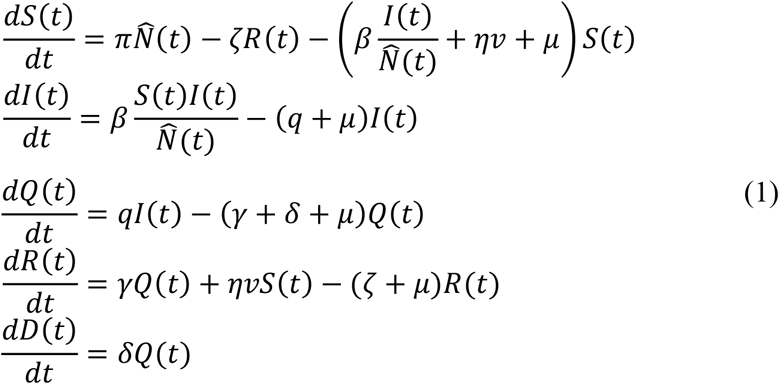

with 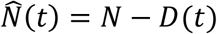 which represents the number of living individuals over time. The population size is assumed to remain constant over time and hence 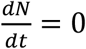 by adjusting *π* = *μ*.

### 3.2 **Multi-Region *SIQRD* Model with Mobility Aspect**

The multi-region SIQRD, denoted by {*SIQRD*}_*k*_, is a generalization of the generic one by simply adding subscript *k* to each compartment, meaning that observation of every state dynamic in *k* regions simultaneously is allowed. The concept of model stratification is commonly used on the analysis of disease spread when it comes to region-related observation (). To capture the effect of massive human mobility on the spread of the virus, the multi-region model accommodates the transfer of individuals from one region to another during the exodus. Several strict assumptions should hold to drop the complexity: only susceptible, inapparent infected, and immune individuals can only get through borders. Infected individuals who develop symptoms will be banned amid borders checking. In contrast, inapparent infected ones are assumed to travel and pass the border checking easily. Human mobility considered in this model is that thrilled by holy days that can drive millions of people to travel from one region to another. This exodus usually takes no longer than a month, and the traveler’s return to their original location is expected to occur. The infections record will be registered in the current place, regardless of the patient’s domicile.

It is undeniable that the travel process, including the type of accommodation, plays a significant role in increasing the chance of infections. However, adding this aspect to the model should increase the complexity and hence will not be considered. Nevertheless, allowing the human transfer from one region to another is still prospective on capturing the effect of the exodus without taking its travel process into account.

Mathematically, the multi-region SIQRD model, which accommodates the inter-region mobility, is given by system (2). The multi-region model generalizes the generic one but adds index *k* and human mobility features.

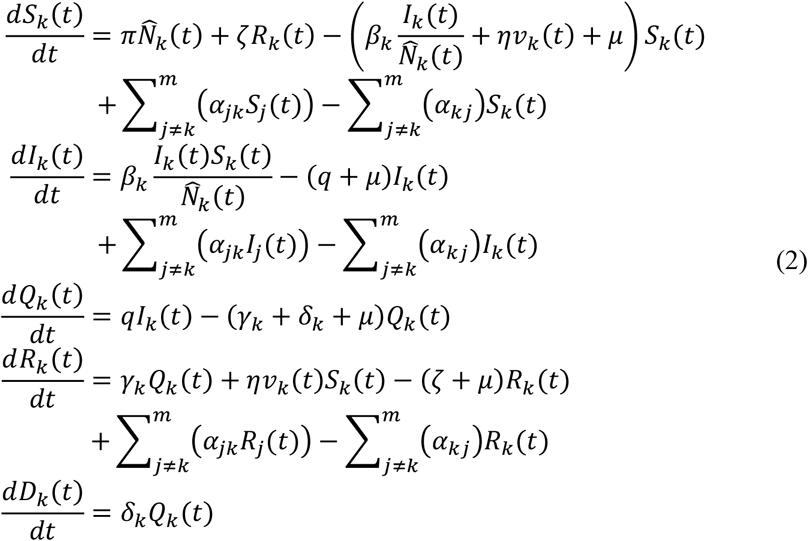

The population size of each region will not be conserved due to the massive mobility from one region to another. However, the sum of all populations in all observed regions will remain constant by assuming no other regions are involved. The initial conditions for variables *Q, R*, and *D* are known from the data, yet that of for *I* is estimated.

According to Table 1, it is clear that some parameters are assumed to depend on regions, meaning its value represents a specific occurrence in each region. The transmission, recovery, and death rate of COVID-19 will depend on the provided data and hence are written with a subscript. The least-square method is used to estimate these parameters by the input of the provided data. Practically, *β, γ, δ*, and *I*_0_ are chosen so that the rendered model to be close to the actual data. However, other parameters are considered more general and hence can be assumed to be uniform for each region.

However, the mobility rate *α*_*jk*_ indicates the share of people traveling from one region to another during the Eid Al-Fitr. Not only be assumed to be a region-specific parameter, but this term is also assumed to vary each time, meaning the mass of mobility will occur occasionally and hence have its dynamic. To be depicted by system (2), the term *α*_*jk*_ affects only the three variables: *S*_*k*_, *I*_*k*_, and *R*_*k*_. The additional term 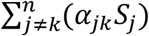 represents the figure of susceptible individuals who enter region k from the rest of the regions. However, term 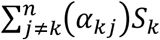 depicts the number of susceptible ones who leave region *k* to other regions. Henceforth, subtracting the two will result in the net change in the susceptible number in the region due to mass exodus. It is noted that the population size in each region may not be conserved, but the sum in all observed regions will remain constant each time.

### 3.3 Method of Estimating the Mobility Rate

According to the constructed model, the mobility aspect is accommodated by parameter *α*_*jk*_, representing the daily portion of people traveling between region *j* and region *k*. Notice that this parameter is time-dependent and can be either positive or negative. Practically, when *α*_*jk*_ is positive, it represents the condition in which portion of people travel from region *j* to region *k*. The greater this value is, the more massive the people travel in the mudik occasion. On the other hand, negative value of *α*_*jk*_ represents the occasion of travel back of people from region *k* to region *j*.

Not only the quantity aspect, the general behavior of *α*_*jk*_ is adjusted to align with the real phenomena: start increasing towards the Eid Al-Fitr and peaking right before the D-Day, followed by the return of people with similar pattern but in reverse. The dynamics are expected to have a significant value near the D-Day of a holy day, making the quantity worth almost zero in other times. By the given assumption, the Dawson Integral Function represents the daily share of the population in a specific region traveling during the exodus.

Based on [31], the general form of the Dawson Integral Function is given as follows:

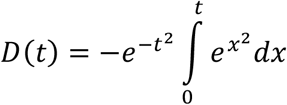

Figure 2(a) depicts the basic graph of Dawson Integral Function equipped with the preferred general behavior: positively increasing and reaching its peak towards the holy day, which represents the mass exodus, and then followed with the reverse pattern in negative values the return. Moreover, when the time of holy day, it is assumed that there is no mobility recorded. However, this function has fixed peaks and amplitude as well as thick tails. In order to align with the expected behavior of mobility rate, the modified Dawson Integral Function is proposed by adding other four additional parameters: *C*_1_, *C*_2_, *t*_0_, and *t*_1_.

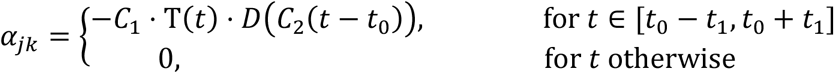

**Figure 2.**
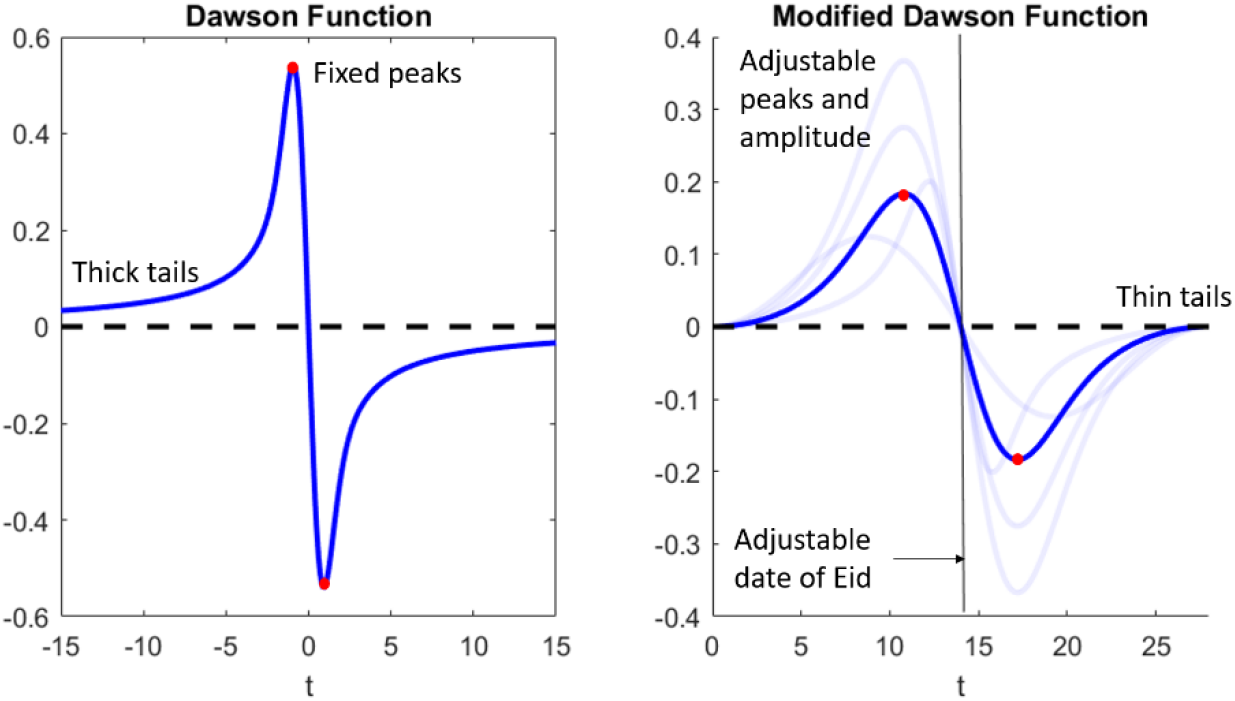
Comparison between the generic Dawson Integral Function with the modified one.

with T(*t*) = (*t* − *t*_0_ + *t*_1_)^2^(*t* − *t*_0_ − *t*_1_)^2^ roles as magnitude adjuster and *D*(*t*) is the Dawson Integral function. By the given modification, several features of the dynamics can be altered:

1. Adjustment of the amplitude: The generic function is multiplied with *C*_1_ so that the amplitude will be as high as *C*_1_.
2. Adjustment of the peaks’ location which is accommodated with adding *C*_2_. On the generic Dawson Function, the location of the peaks is fixed, and thus adding the parameter *C*_2_ will adjust the position of both peaks of the mobility rate.
3. Adjustment of the date of the holy day is accommodated by the adding the parameter *t*_0_. According to the modified Dawson Function, it is clear that when *t* = *t*_0_, then the rate of mobility will be zero. This is relevant due to the assumption in which there is no mobility in the exact day of Eid Al-Fitr.
4. Adding the parameter *t*_1_ will affect the initial time of the mobility. This adjustment is based on the fact that the generic Dawson Function has the thick tail as the time reaches −∞ and ∞. Thus, adding the parameter *t*_1_ to the model can adjust the time in which the mobility is first identified.

Figure 2 compares the general behavior of the generic Dawson Function with the modified one. To be mentioned, the modification yields the flexibility of several features. Depicted by Figure 12(b), both peaks can be adjusted regarding location and amplitude, making it easier to be adjusted to follow the region-specific phenomena. Other than that, both tails are already thin, meaning that mobility is only considered to happen near the holy day.

The complexity is now in estimating all of the additional parameters. Parameter *t*_0_ indicates the occurrence of the holy day, which will be assumed determined based on the initial provided data. Since the data is available from March 27^th^, 2020, then *t*_0_ will be assumed to be *t*_0_ = 407, indicating that the Eid Al-Fitr 2021 will be held 407 days since the first simulation. Other than that, *t*_1_ represents the first day near the holiday which people starts to travel. By assuming that people start to travel in regards to Eid Al-Fitr as14 days before the D-Day, hence *t*_1_ = 14.

On determining the value of *C*_2_, Figure 3 gives insights on how the function change once the value of *C*_2_ is altered. By adjusting the rest of the parameters to be fixed, the change of *C*_2_ is significantly affect the location of both peaks. Increasing the value of *C*_2_ does shift the location of peaks towards the D-Day, and vice versa. This fact indicates that the greater the value is, the closer the peak of the mobility to the holy day. According to [32], it is a commonplace for most Indonesian start to travel 2 days before the holy day, making the mobility peak near this time and packing the road with a long-lasting traffic jam. By the given information, the value of *C*_2_ is determined by forcing that the peaks should occur 2 days prior and after the holy day.

**Figure 3.**
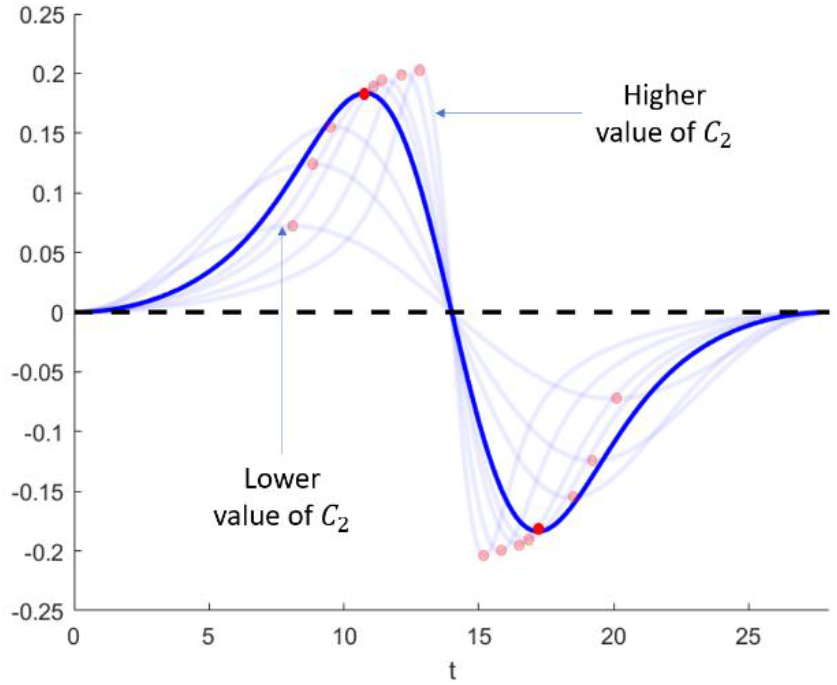
The effect of *C*_2_ change into the location of both peaks.

Lastly, parameter *C*_1_ is expected to be obtained from the provided data since it governs how many people doing the travel. The data used on the estimation is the Origin-Destination (OD) matrix which has been obtained from the phone survey [19]. To be noted that elements consisted in the OD matrix represent the share of population doing the travel due to the holy day. This quantity is a single value and has nothing to do with the daily mobility. The mobility rate *α*_*jk*_, however, depicts the same thing but time dependent. Henceforth, integrating *α*_*jk*_ of time ranging in [*t*_0_ − *t*_1_, *t*_0_] yields the exact representation as that of elements of OD matrix, i.e. share of the certain region’s population traveling from region j to region k. Provided the information for the other three parameters, *C*_1_ can be obtained by forcing that the 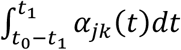 to be close to the data *m*_*jk*_. The detailed information of estimating the *C*_1_ is given in Algorithm 1.

#### Algorithm 1

Estimating the *C*_1_ given the information of OD matrix and other three additional parameters.

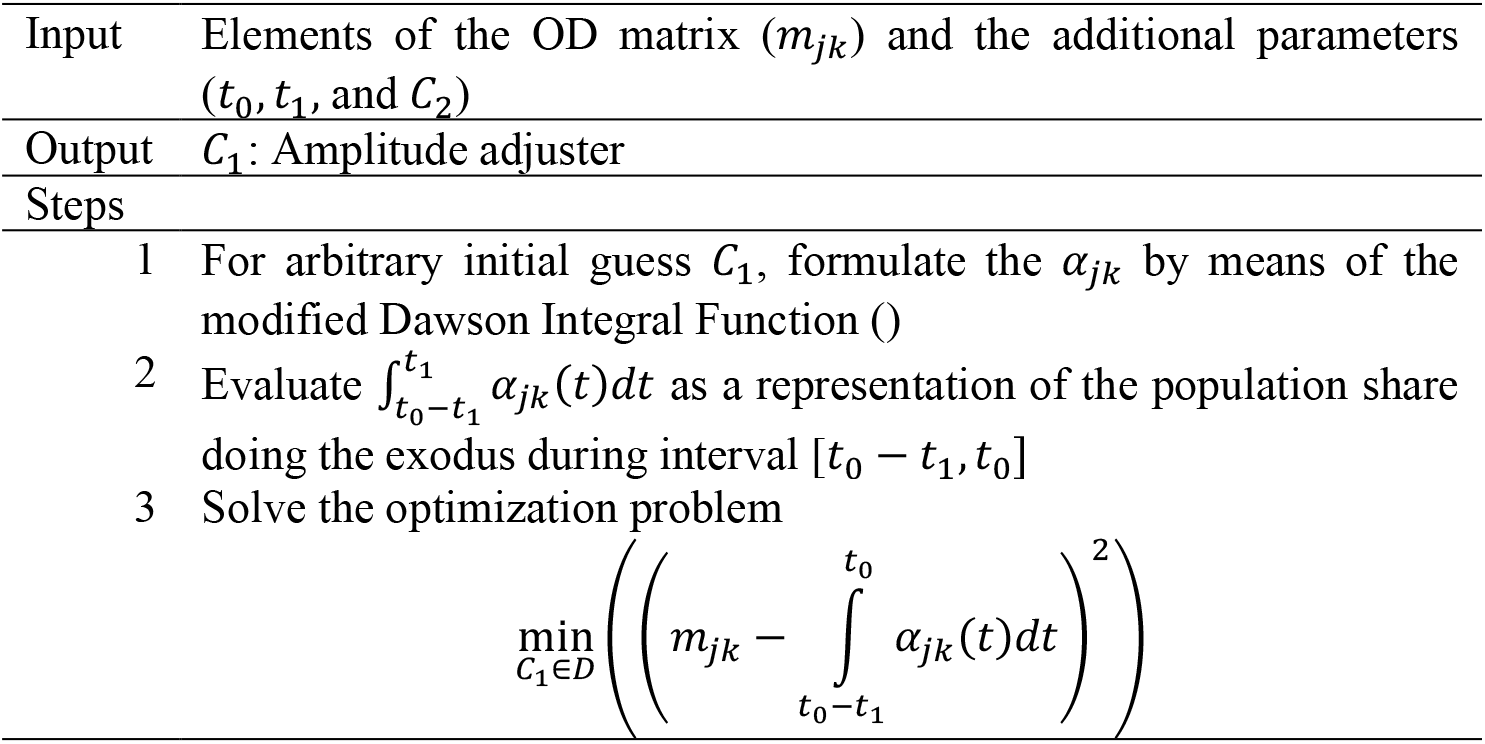

Intuitively, Algorithm 1 decomposes the single value data into time dependent dynamics of mobility rate but conserves its total. For instance, the information of *m*_12_ is being decomposed into *α*_12_(*t*) which has feature of the modified Dawson Integral Function but the total, 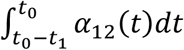 still conform to *m*_12_. Figure 4 shows the visual interpretation of decomposing the data *m*_*jk*_ into time dependent *α*_*jk*_(*t*).

**Figure 4.**
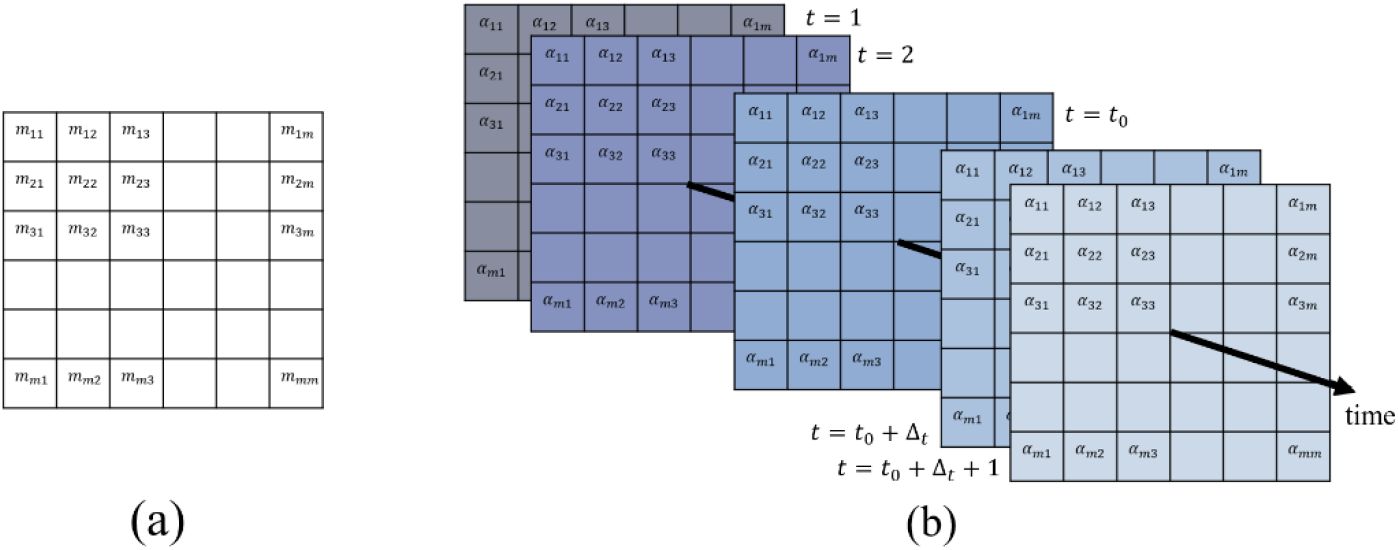
Applying the Algorithm 1 on transforming the OD Matrix data into the time-dependent mobility rate.

## 4 Results

This section provides information related to all parameters’ estimation and the constructed model’s simulation on understanding the effect of the massive mobility on the recorded active cases. According to Table 1, there are parameters in which assumed to be constant and independent of regions. Those mentioned parameters are reinfection rate, quarantine rate, vaccine efficacy, and the natural death and birth rate. Those parameters should be first known by utilizing the existing findings to run the simulations.

According to [30], recovered individuals after being infected can be reinfected by the virus after 1 to 2 years, even though no further research concludes. The quarantine rate is qualified as the non-observable parameters and hence will be assumed to follow the assumption introduced in [33]. The efficacy of vaccines should depend on the vaccine brand being delivered. Since Sinovac is being jabbed at most Indonesians, the vaccine efficacy is assumed to be as high as 57% [34]. The natural death and birth rates are equal and follow the values introduced in [33] to maintain the constant population size.

### 4.1 Parameters’ Estimation

Provided the data of active cases, cumulative recovery, and deaths for all observed regions of Jabodetabek (Jakarta, Bogor, Depok, Tangerang, and Bekasi) and Jabaraya (Jakarta and Bandung Raya) by assuming that the rate of transmission, recovery, and death due to COVID-19 are constant, the single-value estimated parameters are given in Table 2.

**Table 2.** Estimated rate of transmission, recovery, and deaths due to COVID-19 in Jabodetabek and Jabaraya (district/city scale)

| District/City | $\beta$ | $\gamma$ | $\delta$ | $I_0$ |
| --- | --- | --- | --- | --- |
| Jakarta | 0.4106 | 0.1400 | 0.0021 | 2,020 |
| Bekasi, the district | 0.4087 | 0.0474 | 0.0001 | 105 |
| Bogor, the district | 0.4077 | 0.0387 | 0.0001 | 59 |
| Tangerang, the district | 0.4027 | 0.1240 | 0.0028 | 80 |
| Bekasi the city | 0.4082 | 0.0716 | 0.0001 | 189 |
| Bogor, the city | 0.4113 | 0.0525 | 0.0001 | 45 |
| Depok, the city | 0.4116 | 0.0504 | 0.0001 | 149 |
| South Tangerang | 0.4093 | 0.1320 | 0.0032 | 35 |
| Tangerang, the city | 0.4042 | 0.1213 | 0.0018 | 58 |
| West Bandung District | 0.4253 | 0.0900 | 0.0011 | 31 |
| Bandung, the district | 0.3991 | 0.0935 | 0.0001 | 55 |
| Sumedang District | 0.4421 | 0.0485 | 0.0001 | 7 |
| Bandung, the city | 0.3814 | 0.0412 | 0.0001 | 359 |
| Cimahi City | 0.4341 | 0.0140 | 0.0037 | 11 |

### 4.2 Estimated Mobility Rate

According to the phone survey conducted nationally by [19], the results are dominated by respondents from West Java (33.80%), Jakarta (30.95%), Banten (13.07%), Central Java (8.26%), East Java (6.41%), and Yogyakarta (1.61%), and the rest of the share for other provinces in Indonesia. The survey provides information on how many people in the sample intend to travel during the Eid Al-Fitr. By assuming that the sample represents the population, the estimation of the OD matrix can be obtained, as shown in Table 3.

**Table 3.**
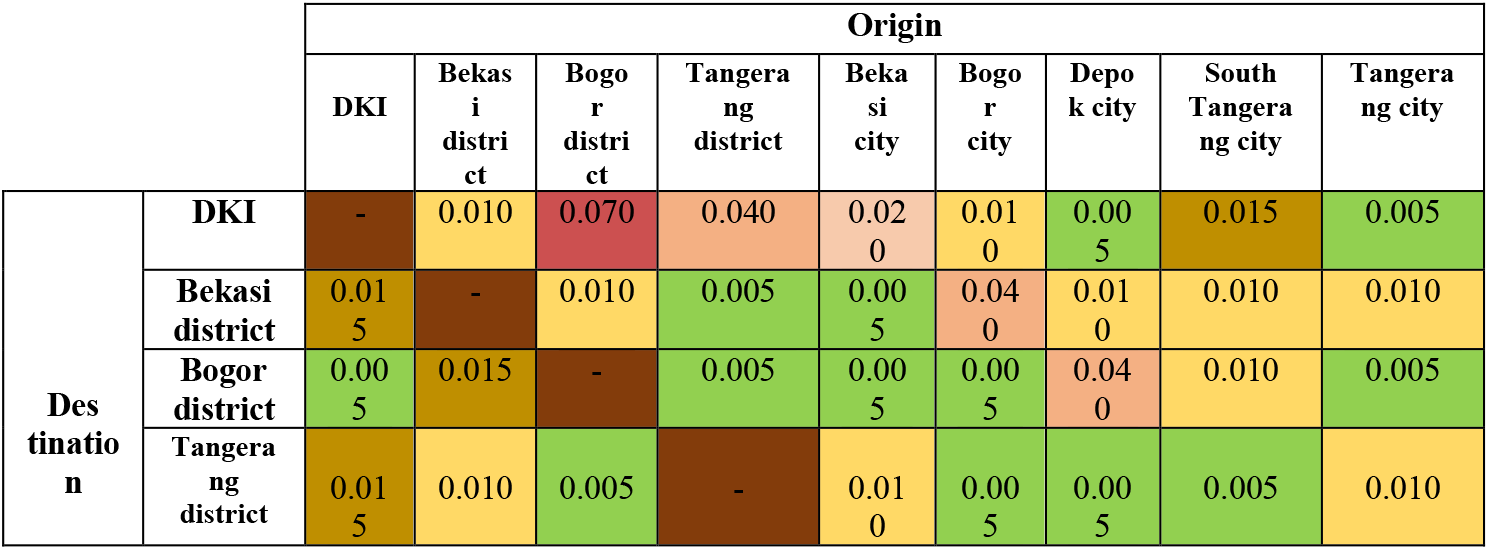
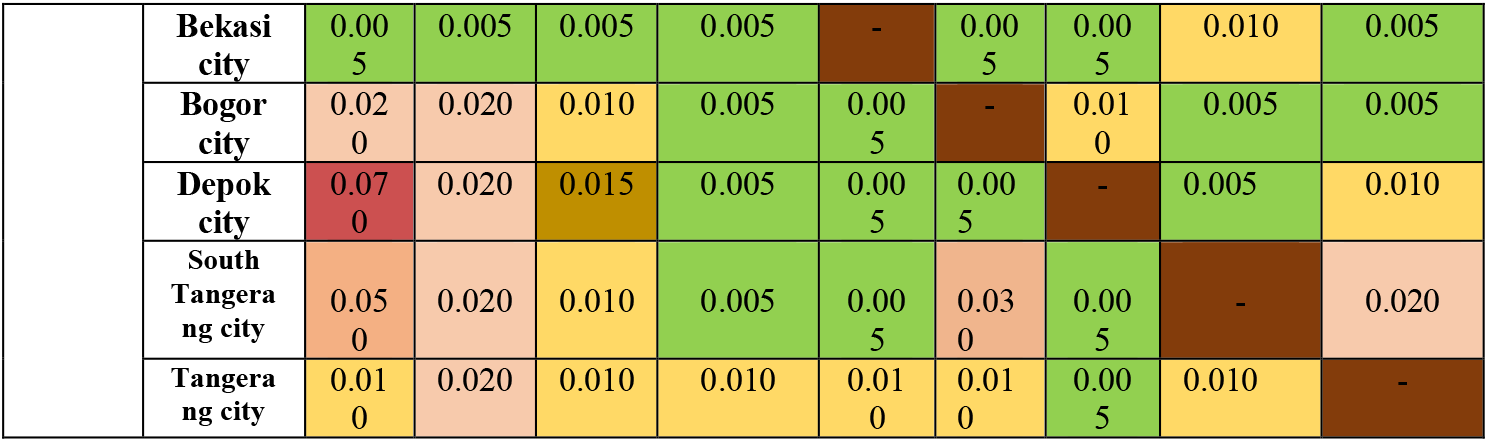
Estimated OD matrix in Jabodetabek (district/city scale).

**Table 4.** Estimated OD matrix in Jabaraya (district/city scale).

|  |  | Origin |  |  |  |  |  |
| --- | --- | --- | --- | --- | --- | --- | --- |
|  |  | DKI | KBB | Bandung district | Cimahi district | Bandung city | Cimahi city |
| Destination | DKI | - | 0 | 0.022 | 0.001 | 0.052 | 0 |
|  | KBB | 0 | - | 0 | 0 | 0.001 | 0 |
|  | Bandung district | 0.042 | 0.021 | - | 0.01 | 0.101 | 0.012 |
|  | Sumedang district | 0.003 | 0.008 | 0.005 | - | 0.012 | 0.005 |
|  | Bandung city | 0.027 | 0.014 | 0.032 | 0.31 | - | 0.01 |
|  | Cimahi city | 0 | 0.021 | 0 | 0.001 | 0.005 | - |

The values originating from Jakarta to West Java are considered to be the highest, which means most mobility depicted from the survey is from Jakarta to West Java. However, some elements are estimated as zeros. This is a direct implication that the values come from the phone survey, which is limited to the recorded sample.

By implementing Algorithm 1, the time-dependent mobility rate having the modified-Dawson-like behavior can be obtained. For visualization, one figure depicting this parameter is shown in Figure 5, representing the mobility rate from Jakarta to Depok City. As expected, mobility’s effect starts to exist 14 days before the D-day and vanishes 14 days later. In the time interval between D-14 until D-Day, the mobility rate is positive, depicting the travel from Jakarta to Depok City. However, after the D-Day, the rate of transmission is negative, representing the return of that traveled.

**Figure 5.**
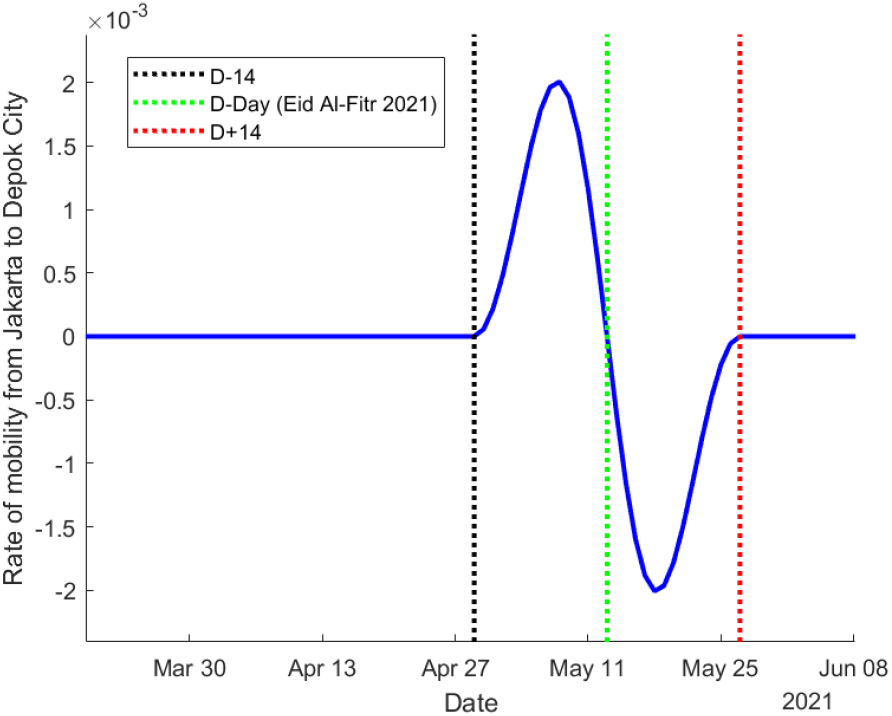
The effect of *C*_2_ change into the location of both peaks

### 4.3 Projected Active Cases with Mobility Aspect

In the previous section, the constant parameters representing the rate of transmission, recovery, and deaths are obtained from the existing data. The obtained parameters are utilized to evaluate the short-term and long-term projected number of active cases of COVID-19. The short-term projection is demonstrated in two scenarios: no exodus or with exodus. Henceforth, the direct effect of massive mobility on the dynamic of active cases can be analyzed.

In Jabodetabek, Figure 6 demonstrates the projected number of active cases in nine cities/districts. That being mentioned, the estimated *β, γ*, and *δ* are assumed to be constant, and hence the model is not able to capture the fluctuating historical. However, the analysis mainly focuses on the projections that occur in 14 days before and after the Eid Al-Fitr (areas between dashed-black and dashed-red lines) in which the mobility rate is significantly non-zero.

**Figure 6.**
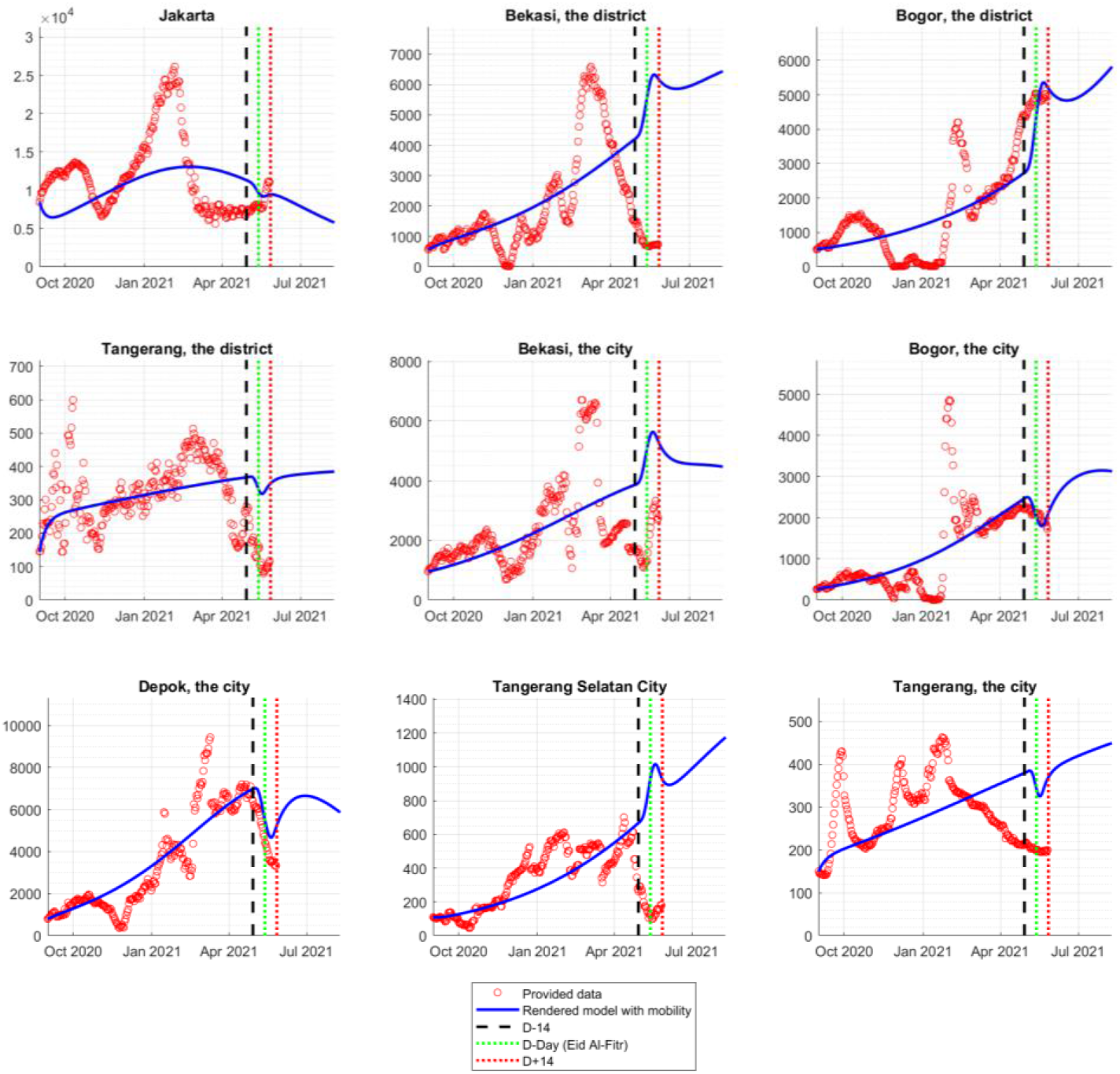
Comparison between the actual data and the rendered model in Jabodetabek during the mass exodus 2021.

In the observed interval, the consideration of the massive mobility directly affects the dynamics of the active case, which can be seen in the interval bracketed by the black and red dashed lines. The four regions, namely Bekasi District, Bogor District, Bekasi City, and Tangerang Selatan City, experienced a significant surge of COVID-19 cases during the mass exodus, while the rest experienced the decline. Specifically, in Depok City, Bogor District, and Bogor City, the rendered model represents the actual data during the mass exodus. In other regions, even though the model seems to overestimate the actual data, the general trend of the actual data can be depicted in the model, except for Bekasi District, Tangerang District, and Tangerang City.

Figure 7 depicts the simulations given for regions of Jabaraya. Overall, the rendered model can predict how the active cases will spike or decline once the OD matrix data is provided. The result for Sumedang District and Cimahi City does underestimate the actual data, but the general behavior is well-captured by the data.

**Figure 7.**
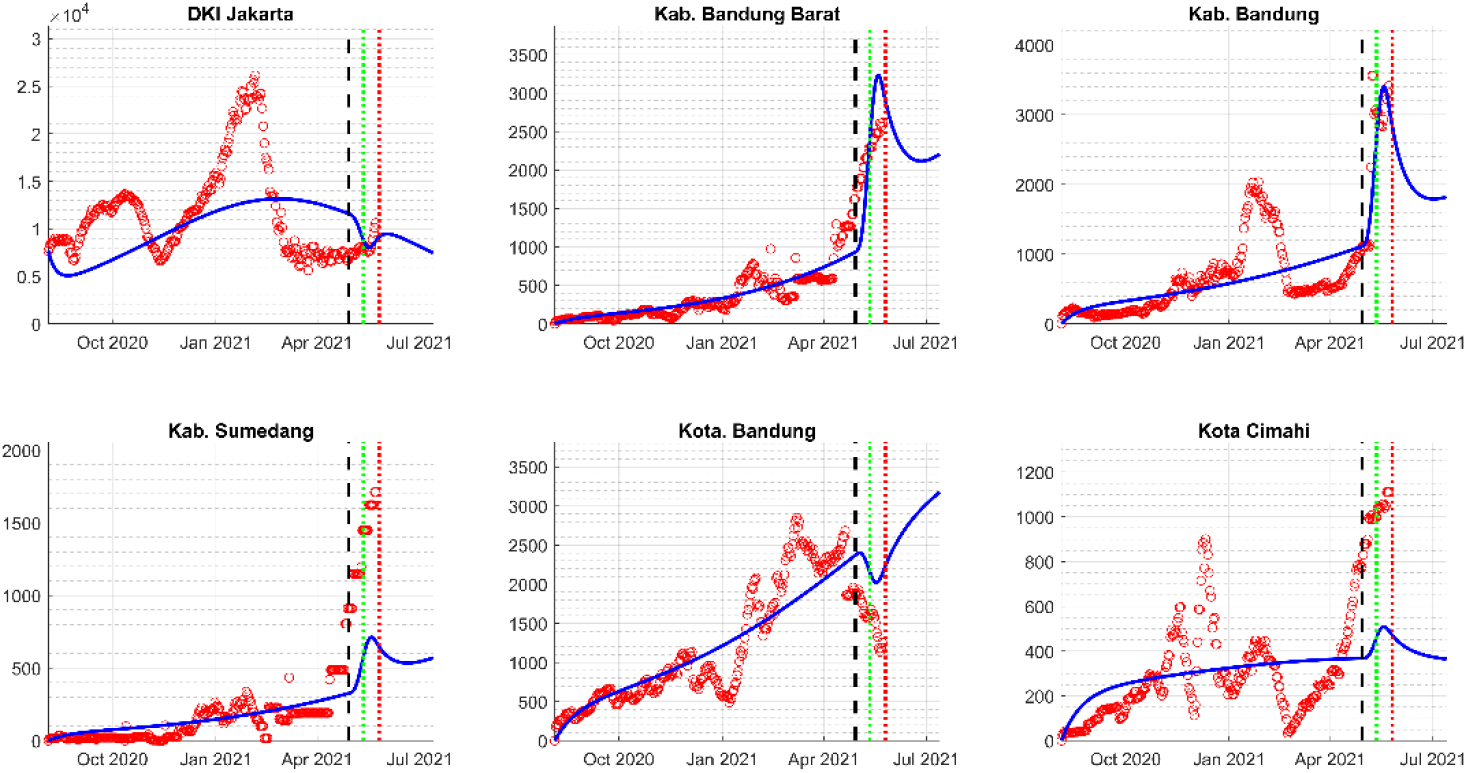
Comparison between the actual data and the rendered model in Jabaraya during the mass exodus 2021.

Additionally, most of the big or agglomerated cities in the observed regions were experiencing a decline in the number of active cases, e.g., Jakarta, Bogor City, Tangerang City, and Bandung City. This result should confirm that most travelers are originating from the big city towards the more rural areas. On the other hand, other regions were experiencing the spike representing the significant supply of susceptible in those regions. Regions with higher spikes tend to be considered as the rural areas which are providing the recreation, e.g. Bandung District and Cimahi City, meaning that not only visiting families and relatives during the holiday, but also visiting some recreational places [35].

## 5 Conclusion

This research has produced a method of estimating the movement of people by representing mobility in the form of a function. The calculation results provide a reasonably good situation compared to the actual data. We use a deterministic model, so in general, the simulation result is the average of the possible occurrences of the given data. The results of this study are considered in formulating policies related to mobility restrictions, especially in urban areas that still have the potential to produce higher cases if the spread cannot be controlled. Although currently, the number of COVID-19 cases in Indonesia is relatively low, the potential for the spread of this disease is still very high, especially with the discovery of new variants. A relaxation that has been massive enough and public awareness to enforce health protocols has also begun to decrease. So one of the efforts that have contributed to helping prevent the spread of COVID-19 is the policy of regulating community mobility, especially in the Java Island area and around the capital city in particular.

## Data Availability

All data produced in the present study are available upon reasonable request to the authors

## Acknowledgement

This research is partly funded by Ministry of Transportation R&D 2021. The authors would like to thank Nunuj Nurdjanah, S.Si, MT., from Litbang Jalan dan Perkeretaapian Indonesia

